# Integrating Imaging-Derived Clinical Endotypes with Plasma Proteomics and External Polygenic Risk Scores Enhances Coronary Microvascular Disease Risk Prediction^†^

**DOI:** 10.1101/2025.08.18.25333844

**Authors:** Rasika Venkatesh, Tess Cherlin, Penn Medicine BioBank, Marylyn D. Ritchie, Marie A. Guerraty, Shefali S. Verma

## Abstract

Coronary microvascular disease (CMVD) is an underdiagnosed but significant contributor to the burden of ischemic heart disease, characterized by angina and myocardial infarction. The development of risk prediction models such as polygenic risk scores (PRS) for CMVD has been limited by a lack of large-scale genome-wide association studies (GWAS). However, there is significant overlap between CMVD and enrollment criteria for coronary artery disease (CAD) GWAS. In this study, we developed CMVD PRS models by selecting variants identified in a CMVD GWAS and applying weights from an external CAD GWAS, using CMVD-associated loci as proxies for the genetic risk. We integrated plasma proteomics, clinical measures from perfusion PET imaging, and PRS to evaluate their contributions to CMVD risk prediction in comprehensive machine and deep learning models. We then developed a novel unsupervised endotyping framework for CMVD from perfusion PET-derived myocardial blood flow data, revealing distinct patient subgroups beyond traditional case-control definitions. This imaging-based stratification substantially improved classification performance alongside plasma proteomics and PRS, achieving AUROCs between 0.65 and 0.73 per class, significantly outperforming binary classifiers and existing clinical models, highlighting the potential of this stratification approach to enable more precise and personalized diagnosis by capturing the underlying heterogeneity of CMVD. This work represents the first application of imaging-based endotyping and the integration of genetic and proteomic data for CMVD risk prediction, establishing a framework for multimodal modeling in complex diseases.

## 1. Introduction

### 1.1. CMVD contributes to the large burden of ischemic heart disease

Coronary microvascular disease (CMVD) is increasingly recognized as a major contributor to the global burden of ischemic heart disease (IHD)^1–3^. Unlike obstructive coronary artery disease (CAD), which involves large-vessel atherosclerosis, CMVD affects the smaller vessels of the heart and can present clinically in patients as angina, ischemia, myocardial infarction with no obstructed coronary arteries, or increased major adverse cardiovascular events^4^. Studies estimate that CMVD may account for 30–50% of cases of angina in patients without obstructive CAD, underscoring its critical role in IHD^1,3^. CMVD is often undetected through standard cardiac imaging methods, leading to underdiagnosis; advanced techniques such as perfusion positron emission tomography (PET) currently represent the non-invasive gold standard for diagnosis^5^. Despite its high prevalence, no targeted therapies for CMVD currently exist, and its diagnosis relies on specialized testing that limits routine clinical use^5,6^. The diagnostic challenges and lack of targeted treatment for CMVD reflect a limited understanding of its pathophysiology^7^. CMVD involves both structural and functional changes to microvasculature, driven by cardiovascular comorbidities^1,5^. The genetic and molecular underpinnings of the condition remain poorly understood, resulting in underdiagnosis and insufficient treatment. Improved molecular characterization incorporating advanced imaging, genetics, and proteomics holds promise for better risk stratification, improved diagnostic approaches, and eventual therapeutic development for this condition.

### 1.2. Pathophysiological variation in CMVD supports the presence of endotypes

CMVD is a highly heterogeneous disease with variable clinical presentation^7^. Existing clinical studies rely on measurements of coronary microvascular function, using invasive and non-invasive imaging tools to measure myocardial blood flow and flow reserve (MBFR). Reduced MBFR suggests impaired microvascular function and has been associated with increased mortality^2,8^. Clinical studies using cardiac perfusion PET imaging have shown that impaired MBFR can result from either increased resting flow or blunting of stress flow, indicating that multiple pathophysiologic pathways may exist, reflecting biologically distinct disease subtypes^8,9^. These disparate clinical mechanisms suggest the presence of different endotypes of CMVD manifesting with distinct flow patterns. A one-size-fits-all approach to modeling CMVD is insufficient for precise diagnosis and management. We stratified patients into endotypes in this study based on image-derived flow parameters. This approach enables more nuanced modeling of intermediate and heterogeneous cases, laying the groundwork for precision medicine in CMVD.

### 1.3. Existing early risk prediction methods for CMVD are limited

Current diagnostic approaches for CMVD rely on clinical measurements derived from perfusion PET imaging, alongside tracking of clinical risk factors like cholesterol levels and comorbidities such as diabetes as they emerge. As a result, the condition is often underdiagnosed, and patients can be misclassified. Biomarker-based risk prediction incorporating genetic and proteomic features could provide an alternative for identifying individuals at risk for CMVD prior to the emergence of clinical risk factors. However, the development of predictive models for CMVD has been limited by the lack of large-scale genome-wide association studies (GWAS), and to date, no polygenic risk scores (PRS) have been developed for the disease. The clinical model developed by Prescott *et al*. is the only existing risk prediction model for CMVD. This study incorporated integrated proteomics and achieved relatively strong performance (AUROC = 0.61-0.66) but still relies on lab measurements and clinical variables that develop over time, such as diabetes and hypertension^10^.

CAD GWAS have historically enrolled heterogeneous IHD populations that include both obstructive CAD and CMVD, making CAD GWAS effectively IHD GWAS^3^. Despite their clinical differences, CMVD and obstructive CAD may arise from overlapping genetic mechanisms, including those involved in inflammation and microvascular remodeling^3^. While many CAD-associated loci are relevant to CMVD, our previous work has shown that CMVD also involves distinct loci not captured by CAD studies^11^. This overlap and divergence indicate that CMVD shares components of the broader IHD genetic architecture while maintaining unique molecular underpinnings^3,6,11^. In this study, we evaluate the utility of an externally derived PRS for coronary artery disease (CAD), a related phenotype with overlapping enrollment criteria, to assess the potential for cross-phenotype genetic risk modeling. We present the first machine learning framework for CMVD risk prediction that integrates genomic and proteomic data without the need for clinical input. By incorporating endotype-based stratification from imaging, we also demonstrate improved classification performance. We demonstrate the ability to predict risk earlier than clinical features emerge across traditional and intermediate groups, thereby greatly enhancing the clinical utility of the model for patient prioritization and potential for new diagnostic approaches.

## 2. Methods

### 2.1. PennMedicine BioBank Study Population

The Penn Medicine BioBank (PMBB) represents a diverse population of 57,170 participants who have been genotyped and imputed using the TOPMed reference panel (Version r2 2020)^12,13^. Each participant’s genetic data is linked with their Electronic Health Record (EHR), which includes perfusion PET stress testing data. We identified 4,007 patients who underwent Rubidium-82 perfusion PET stress testing at the Hospital of the University of Pennsylvania as part of routine clinical care and enrolled in PMBB. As shown in **Table 1**, 829 of these participants had high-quality perfusion PET imaging, genetic, and proteomics data (92 proteins from OLINK Cardiovascular II panel) available and were included in the final analysis. This comprises one of the largest multi-omics datasets for CMVD, with integrated imaging, genetics, and proteomics data.

**Table 1.**
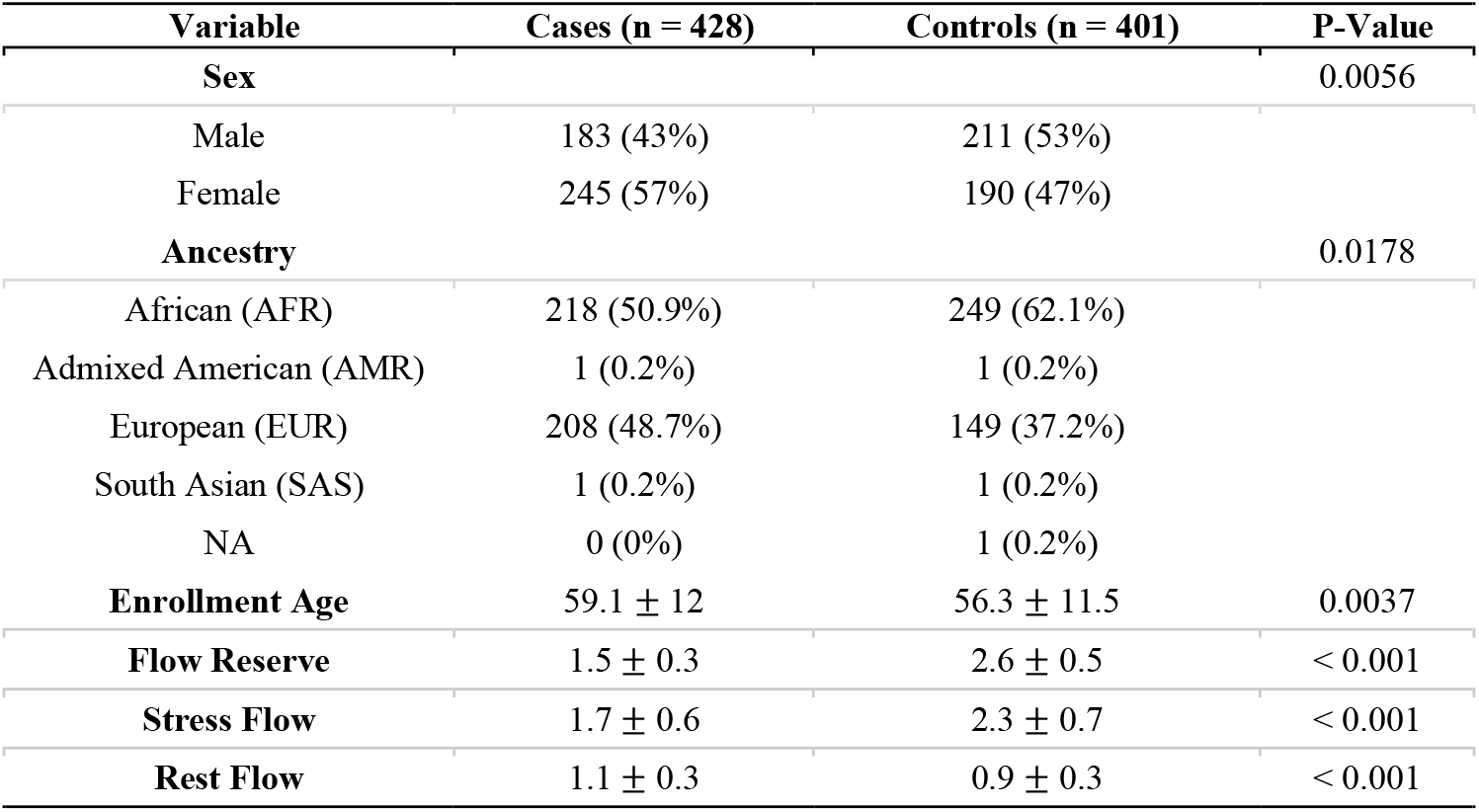
Demographic and clinical information for the study population. P-values calculated using Pearson’s Chi-Squared Test (Sex, Ancestry) and Welch’s T-Test (Age, Imaging)

| Variable | Cases (n = 428) | Controls (n = 401) | P-Value |
| --- | --- | --- | --- |
| **Sex** |  |  | 0.0056 |
| Male | 183 (43%) | 211 (53%) |  |
| Female | 245 (57%) | 190 (47%) |  |
| **Ancestry** |  |  | 0.0178 |
| African (AFR) | 218 (50.9%) | 249 (62.1%) |  |
| Admixed American (AMR) | 1 (0.2%) | 1 (0.2%) |  |
| European (EUR) | 208 (48.7%) | 149 (37.2%) |  |
| South Asian (SAS) | 1 (0.2%) | 1 (0.2%) |  |
| NA | 0 (0%) | 1 (0.2%) |  |
| **Enrollment Age** | 59.1 ± 12 | 56.3 ± 11.5 | 0.0037 |
| **Flow Reserve** | 1.5 ± 0.3 | 2.6 ± 0.5 | < 0.001 |
| **Stress Flow** | 1.7 ± 0.6 | 2.3 ± 0.7 | < 0.001 |
| **Rest Flow** | 1.1 ± 0.3 | 0.9 ± 0.3 | < 0.001 |

All patients received dipyridamole or regadenoson for coronary vasodilation, and the imaging data was analyzed using Siemens Syngo MBF or Invia Corridor 4DM. Patients with incomplete or poor-quality myocardial blood flow reserve (MBFR) or proteomic data were excluded from the analysis during quality control, resulting in the final cohort size of 829 participants. The proteomics data for these patients were normalized using intensity normalization and subsequently log_2_ transformed. CMVD case-control status was defined by MBFR, with cases exhibiting MBFR < 2, indicative of impaired coronary microvascular function, and controls having MBFR ≥ 2, indicative of normal function^8^. Demographic covariates of age, sex, and principal components (PCs) accounting for population stratification based on genetic ancestry (determined in Verma *et al*.^12^) were selected based on statistical significance to CMVD case-control status (**Table 1)**.

### 2.2. Polygenic Risk Score Calculation

To investigate the genetic contribution to CMVD risk, we calculated external PRS using the --score function in PLINK, based on previously published GWAS summary statistics from Aragam *et al*., a large-scale study of CAD conducted on 1,165,690 participants^14–16^. This PRS aggregates the effects of genetic variants associated with CAD risk and applies them to our samples to estimate each individual’s inherited predisposition to vascular disease^16,17^. The variants identified in CAD GWAS, particularly those relevant to CMVD, provide a biologically informed basis for constructing PRS to assess shared susceptibility.

We constructed 3 PRS models: 1) CAD PRS: using 235 genome-wide significant SNPs identified in the CAD GWAS that also had genotypes in our PMBB participants, 2) CMVD Targeted PRS: 15 SNPs previously reported as significantly associated with CMVD in a targeted association study from CAD GWAS summary statistics^11^ and 3) CMVD Targeted + GWAS PRS: combining the 15 SNPs with 5 SNPs for *CAPN2* (rs6700366), *ERC1* (rs115240131), *PLA2G5* (rs11573191), *AADAC* (rs1969348), and *ADK* (rs839696) identified as significant via a prior GWAS, fine-mapping, and TWAS conducted specifically for CMVD^11^. The list of SNPs included in each PRS is presented in **Supplemental Table** S**1**. This modeling approach enabled us to compare the predictive utility of broadly CAD-related SNPs in creating CMVD-focused genetic risk profiles in our cohort. The PRS was fitted with a LASSO regression, using case/control status after splitting the dataset into a 80% training and 20% testing split across 100 random iterations, balanced by the case/control ratio^18–20^. Model performance was evaluated across metrics, including the area under the receiving operator characteristic (AUROC), precision-recall curve (AUPRC), F1-score, balanced accuracy (BA), and R^2^ calculated using the Matthews Correlation Coefficient (MCC)^21,22^.

### 2.3. CMVD Disease State Endotyping

We applied unsupervised k-means clustering to an independent cohort of 3,130 PMBB individuals with recorded global reserve, rest, and stress myocardial blood flow values to identify distinct physiological endotypes associated with CMVD^8,23^. The distribution of these imaging measurements is shown in **Supplemental Figure S1**. Prior to clustering, all variables were standardized and outliers removed to ensure comparability. The optimal number of clusters was determined using the silhouette score and the elbow method^24^. After defining CMVD endotypes in this cohort, we projected our independent cohort of 829 individuals with OLINK proteomics data onto the identified clusters by calculating the Euclidean distance between each proteomics individual’s reserve, rest, and stress values and the centroids of the defined clusters, assigning each individual to the closest endotype^8,24^.

### 2.4. Risk Modeling

#### 2.4.1. Model Architectures and Feature Sets

To investigate the predictive value of genetic and proteomic data for CMVD risk prediction, we developed and compared several machine learning models across a range of feature inputs. We trained 4 distinct linear and nonlinear model types using Scikit-learn and Pytorch: logistic regression with elastic net regularization (EN), eXtreme gradient boosting (XGBoost), deep autoencoders, and fully connected feedforward neural networks (FNN)^25,26^. Each modeling approach was applied to a standardized set of 8 input feature types, described in **Table 2**, to systematically assess performance and interpretability.

**Table 2.**
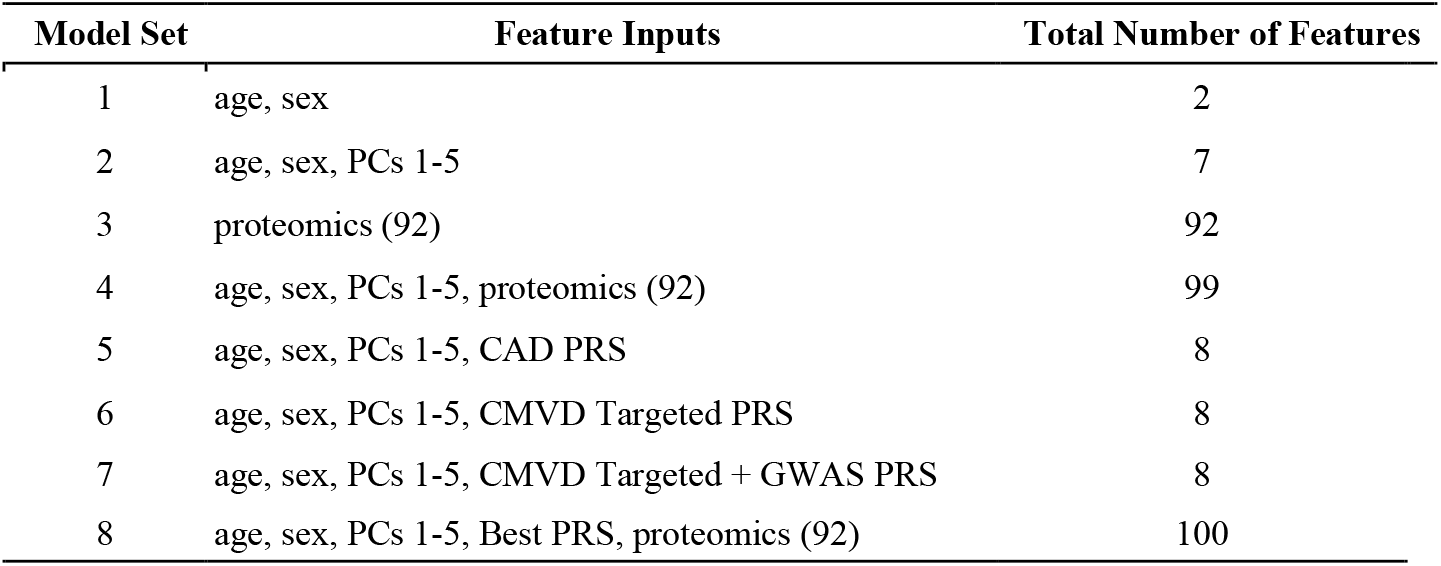
Model feature inputs and total number of features.

| Model Set | Feature Inputs | Total Number of Features |
| --- | --- | --- |
| 1 | age, sex | 2 |
| 2 | age, sex, PCs 1-5 | 7 |
| 3 | proteomics (92) | 92 |
| 4 | age, sex, PCs 1-5, proteomics (92) | 99 |
| 5 | age, sex, PCs 1-5, CAD PRS | 8 |
| 6 | age, sex, PCs 1-5, CMVD Targeted PRS | 8 |
| 7 | age, sex, PCs 1-5, CMVD Targeted + GWAS PRS | 8 |
| 8 | age, sex, PCs 1-5, Best PRS, proteomics (92) | 100 |

Model 1 used only age at enrollment and biological sex extracted from EHR as demographic features. Model 2 accounted for population structure by including the top 5 PCs as well as age and sex. Models 3 and 4 incorporated 92 plasma proteins, resulting in 92 and 99 features, respectively. Models 5-7 incorporated the 3 different PRS, each using 8 features with age, sex, and PCs 1-5. Model 8 combined the best-performing PRS with proteomics (100 features) and was extended to a multiclass prediction task based on CMVD subtype clustering results. All models underwent hyperparameter tuning and top feature were assessed. We employed this modeling design to assess model performance and biologically-informed feature contributions comprehensively.

#### 2.4.2. Elastic-Net Logistic Regression (EN)

To classify CMVD risk status, we implemented a logistic regression model with Elastic Net regularization (EN) to handle high-dimensional and potentially correlated input features^27^. After separating the data into 80% training and 20% test splits, the EN model was trained with nested cross-validation for hyperparameter tuning. We tested a range of inverse regularization strengths (Cs=10) across multiple parameters (L1 ratios from 0.1 to 1.0), using 5-fold cross-validation and AUROC as the optimization metric^27^. To ensure robustness, the entire training and evaluation process was repeated across 100 stratified and Z-score normalized training and testing splits. Models were evaluated using AUROC, AUPRC, F1-score, BA, precision, recall, and Brier score^28,29^. Feature importance was assessed using SHapley Additive exPlanations (SHAP) values for local additive explanations of the model output from a LinearExplainer^30^.

#### 2.4.3. eXtreme Gradient Boosted Trees (XGBoost)

To capture nonlinear interactions among the features, we implemented a gradient boosting framework using XGBoost for binary classification of CMVD status^31^. For each of 10 independent iterations, the data were stratified by CMVD status and split into training (80%) and test (20%) sets, with a further 20% validation set split from the training data. Hyperparameter tuning was performed on the validation set using Optuna with 10 trials per iteration, leveraging a tree-structured Parzen estimator (TPE) sampler^32^. Tuned parameters included tree depth, learning rate, subsample, and column sampling ratios, L1/L2 regularization (alpha, lambda), minimum loss reduction (gamma), and scaled positive weight for class imbalance. Each model was trained with the binary logistic objective and early stopping based on validation AUROC, with a patience of 20 rounds^33,34^. The best model per iteration was selected and evaluated on the held-out test set. Model performance was assessed across metrics including AUROC, AUPRC, F1-score, precision, recall, BA, and Brier score^28,29^. Feature importance was evaluated SHAP values from TreeExplainer for the best-performing model to interpret the contributions of top features^30,35^.

#### 2.4.4. Autoencoder

A deep autoencoder architecture was also employed to attempt to analyze the high-dimensional protein expression profiles for CMVD detection^36^. In each iteration, the dataset was randomly divided into training and testing sets with an 80% and 20% split, stratified by CMVD. The training set was further split into 20% validation subsets for hyperparameter optimization. The encoder consists of multiple fully connected layers, with the number of layers and the number of units per layer optimized via hyperparameter tuning. Each encoder layer applied a linear transformation followed by batch normalization, ReLU activation, and dropout to prevent overfitting^37^. The decoder then reconstructed the original input from the compressed latent representation. A classification head was attached to the final layer to predict CMVD using a sigmoid activation function^37^. This architecture enabled simultaneous learning of a compressed representation and a classification output from the same latent features. The autoencoder was trained using the Adam optimizer, with the learning rate selected from a log-uniform distribution between 1E-04 and 1E-02^38^.

The model minimized a combined loss comprising mean squared error (MSE) for input reconstruction and binary cross-entropy (BCE) for classification, ensuring the encoded features were representative of the protein profiles and predictive of CMVD status^39^. We employed early stopping with a patience of 5 epochs, based on the combined validation loss, and monitored it over a maximum of 300 epochs^36^. A batch size of 64 was used and poorly performing trials were pruned during hyperparameter tuning. Model performance was assessed through repeated stratified splits and evaluation over 10 iterations. Performance metrics were calculated on the independent test sets and averaged across iterations to ensure robustness. Metrics included the AUROC, AUPRC, precision, recall, F1-score, BA, and Brier score. SHAP values were computed using DeepExplainer on the best model to interpret feature contributions to the CMVD classification^30^.

#### 2.4.5. Feedforward Neural Network (FNN)

A feedforward neural network (FNN) was developed to model binary classification for CMVD risk^40^. In each iteration, the dataset was randomly divided into training and testing sets with an 80% and 20% split, stratified by CMVD status to maintain class balance. The training set was further split into 20% validation subsets for hyperparameter optimization. Input features were standardized using z-score normalization. The network architecture consisted of 2 or 3 fully connected hidden layers, selected based on hyperparameter tuning per iteration, followed by batch normalization, ReLU activation, and dropout regularization to enhance convergence stability and reduce overfitting. The size of each layer, dropout rates, and learning rate were also optimized using Optuna with a randomized search^41^. The final output layer used a single sigmoid-activation function, producing probabilistic outputs for CMVD risk. BCE was used as the loss function, and training was performed with the Adam optimizer. Early stopping was applied based on validation loss with a fixed patience threshold of 5 epochs to minimize overfitting^38,40^.

To ensure robustness and generalizability, the training pipeline was repeated across 10 random stratified splits. For each iteration, the model was retrained using the best hyperparameters identified via nested optimization. Performance metrics—including AUROC, AUPRC, F1-score, BA, precision, recall, and Brier score—were calculated on held-out test data and summarized using means and standard deviations. Model explainability was assessed using SHAP, applied to the best-performing iteration to identify the most influential features driving CMVD risk prediction. SHAP values were computed using KernelExplainer on a reduced background set, and results were visualized through summary plots and ranked feature importance^30^.

#### 2.4.6. Multi-Class Classification

To predict CMVD endotypes identified through unsupervised clustering of perfusion PET imaging data, we implemented a supervised multiclass classification framework using both XGBoost and multinomial logistic regression (MLR)^42^. The input data consisted of integrated transcriptomic and proteomic features, with clustering-derived labels representing distinct CMVD endotypes. For model training and evaluation, the dataset was randomly split into 80% training and 20% testing subsets in each iteration, with stratification by endotype label to preserve class distributions. The training set was further divided into 80% training and 20% validation subsets.

We performed 10 independent iterations of model training for both XGBoost and MLR to evaluate performance stability across data splits. For XGBoost, Optuna optimization was employed within each iteration to tune hyperparameters over 10 trials, aiming to minimize multiclass log-loss on the validation set. Parameters included tree depth, learning rate, number of estimators, L1/L2 regularization, and feature and sample subsampling rates^31^. For MLR, we optimized regularization strength, penalty type, solver, maximum iterations, tolerance, and L1 ratio (for EN)^27^. Final models in each iteration were trained with early stopping and evaluated on the held-out test set. Metrics collected per iteration included macro-averaged precision, recall, F1-score, AUROC, AUPRC (one-vs-rest), and BA. Model interpretability was assessed using SHAP values from TreeExplainer to identify the most influential features contributing to each endotype classification^43^.

## 3. Results

### 3.1. PRS Performance

PRS models were evaluated for their ability to predict binary CMVD case-control status using different sets of SNPs, as seen in **Table 3**. Across all SNP sets, model performance improved with the inclusion of age, sex, and genetic ancestry covariates (PCs 1–5), as reflected in higher AUROC, AUPRC, and R^2^ values. The CMVD-targeted PRS, using 15 CMVD-related SNPs, demonstrated improved predictive performance compared to the CAD PRS. Incorporating the additional 5 SNPs from CMVD GWAS also showed improvement over the CAD PRS, achieving similar performance to the targeted approach. When combined with covariates, the CMVD-targeted SNP models achieved the highest AUROC (0.601) and R^2^ (0.142), indicating that focusing on CMVD-relevant genetic variants when using weights from a related phenotype enhances performance. These findings highlight the utility of targeted PRS approaches in improving genetic risk prediction.

**Table 3.** PRS model performance in CMVD risk prediction. Metrics reported as mean (standard deviation).

| PRS Model | Feature Inputs | AUROC | AUPRC | BA | F1-Score | $R^2$ |
| --- | --- | --- | --- | --- | --- | --- |
| CAD | PRS | 0.569 (0.033) | 0.590 (0.040) | 0.543 (0.030) | 0.544 (0.068) | 0.086 (0.060) |
| CAD | PRS, age, sex, PCs | 0.587 (0.031) | 0.595 (0.040) | 0.554 (0.027) | 0.552 (0.035) | 0.108 (0.054) |
| CMVD Targeted | PRS | 0.572 (0.029) | 0.570 (0.035) | 0.546 (0.025) | **0.624 (0.035)** | 0.106 (0.058) |
| CMVD Targeted | PRS, age, sex, PCs | **0.601 (0.028)** | **0.598 (0.036)** | **0.571 (0.024)** | 0.569 (0.031) | **0.142 (0.048)** |
| CMVD Targeted + GWAS | PRS | 0.543 (0.025) | 0.555 (0.035) | 0.521 (0.021) | 0.540 (0.033) | 0.042 (0.033) |
| CMVD Targeted + GWAS | PRS, age, sex, PCs | 0.595 (0.028) | 0.597 (0.035) | 0.570 (0.025) | 0.588 (0.032) | 0.141 (0.050) |

### 3.2. Binary Risk Prediction Model Performance

We evaluated the performance of 8 predictive models incorporating demographic variables (age, sex), genetic ancestry (PCs), PRS, and plasma proteomics using multiple performance metrics, including the AUROC, AUPRC, BA, and F1-score. **Figure 1** summarizes the comparative performance across models. Full results across all metrics are provided in **Supplemental Table S2**. EN and XGBoost consistently outperformed other models across evaluation metrics, particularly when proteomic features were included alongside demographic and genetic variables. With only age and sex as inputs, model discrimination was modest (AUROC = 0.57), but when proteomics were added in Models 3, 4, and 8, AUROC values rose substantially. The EN improved from 0.577 to 0.675 in Model 3, and XGBoost from 0.573 to 0.659. These trends held when including PCs and PRS, further reinforcing the additive predictive value of proteomic data. EN achieved the best overall performance across AUROC (0.676), AUPRC (0.683), and Brier Score (0.229) when using the feature set of age, sex, proteomics, and PCs (Model 4). XGBoost excelled in recall, reaching a high of 0.966 with the CMVD Targeted PRS and 0.907 with the full proteomic panel, indicating that it is particularly effective in identifying true positives. While the autoencoder and feedforward neural network (FNN) models performed more modestly, they still benefited from proteomic inclusion, demonstrating significant increases in performance. SHAP feature importances for all models are reported in **Supplemental Table S3** and SHAP summary plots for the top models (Models 3, 4, and 8) are available in **Supplemental Figures S2-4**. Overall, these results underscore the added value of proteomics in capturing biological signals relevant to CMVD and highlight the strengths of EN and XGBoost in classification performance.

**Fig. 1.**
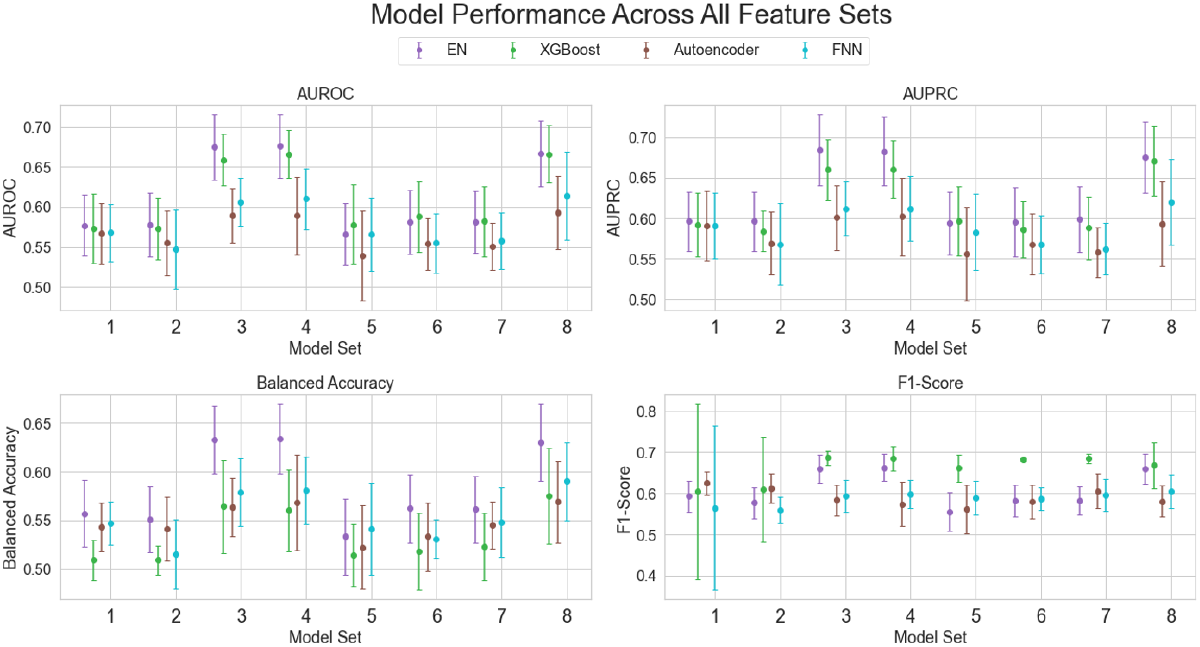
Model performance (AUROC, AUPRC, BA, F1-score) across all feature sets for EN, XGBoost, Autoencoder, and FNN models.

### 3.3. CMVD Endotype Clustering and Multiclass Prediction

The k-means clustering analysis identified distinct endotypes based on global reserve, rest, and stress values, with an overall inertia of 1035.87, indicating meaningful separation between clusters. The elbow plot for determining the optimal number of clusters is shown in **Supplemental Figure S5**; the data are best represented by 4 clusters, as shown in **Figure 2**^24^. Class 0 (n = 290) represented the classic CMVD cases, characterized by reduced flow reserve and low stress flow values, consistent with impaired microvascular function. In contrast, Class 2 (n = 110) comprised the classic control group, with the highest reserve values. Class 1 (n = 233) and Class 3 (n = 192) reflect intermediate groups, with Class 3 characterized by low or borderline reserve, likely due to elevated resting flow. Together, these clusters define a spectrum of microvascular phenotypes ranging from healthy controls to individuals with established CMVD and intermediate physiological profiles^8^.

**Fig. 2.**
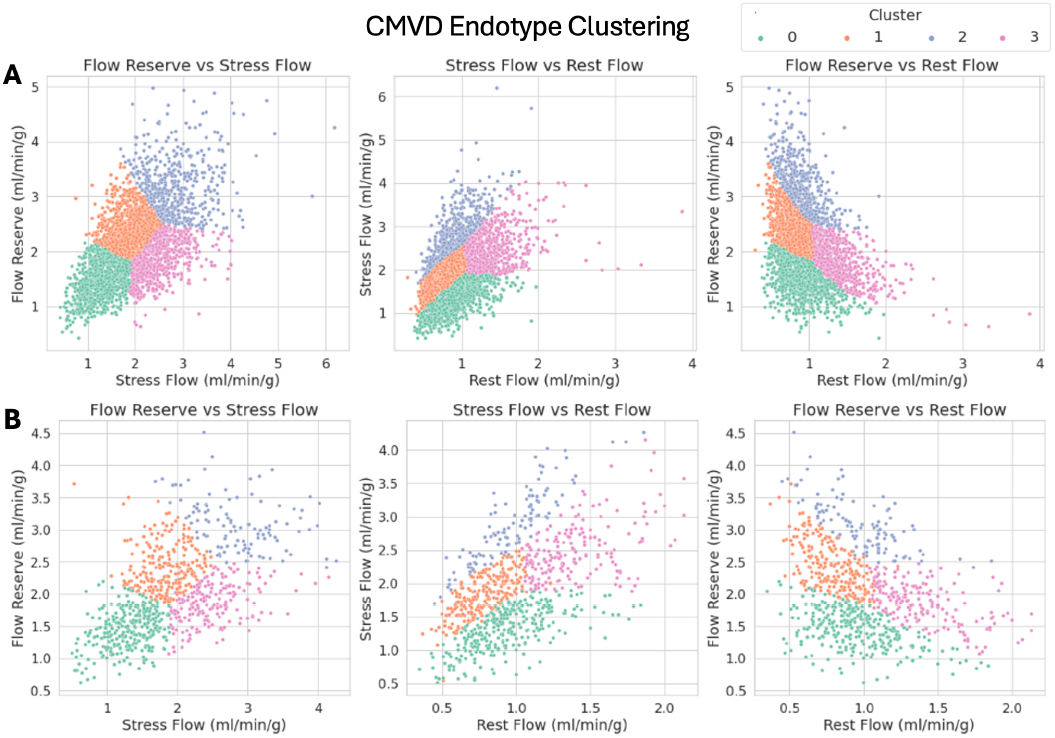
CMVD endotype clustering by flow stress, rest, and reserve. A) Unsupervised clustering of PMBB individuals not in OLINK. B) Projection of 829 OLINK individuals onto defined clusters.

To further characterize these endotypes, we trained XGBoost and MLR multiclass classifiers using PRS and proteomics, with model interpretation performed using SHAP analysis. The XGBoost multiclass model showed substantial improvement in performance relative to the binary classification approaches, as shown in **Table 4**.

**Table 4.** Performance metrics for multiclass model (mean, standard deviation) reported for each class and overall performance.

| Class | AUROC | AUPRC | Precision | Recall | BA | F1-Score | Brier Score |
| --- | --- | --- | --- | --- | --- | --- | --- |
| Class 0 | 0.723 (0.036) | 0.584 (0.065) | 0.491 (0.033) | 0.712 (0.053) | 0.580 (0.030) | 0.656 (0.030) | 0.197 (0.009) |
| Class 1 | 0.648 (0.048) | 0.434 (0.056) | 0.419 (0.067) | 0.389 (0.072) | 0.402 (0.063) | 0.587 (0.042) | 0.192 (0.006) |
| Class 2 | 0.721 (0.054) | 0.345 (0.084) | 0.389 (0.173) | 0.145 (0.067) | 0.206 (0.094) | 0.553 (0.038) | 0.107 (0.005) |
| Class 3 | 0.669 (0.028) | 0.389 (0.047) | 0.394 (0.045) | 0.295 (0.094) | 0.333 (0.075) | 0.580 (0.037) | 0.168 (0.005) |
| All Classes | 0.690 (0.024) | 0.438 (0.030) | 0.434 (0.041) | 0.447 (0.031) | 0.422 (0.039) | 0.447 (0.031) | 0.166 (0.004) |

While the binary models achieved AUROCs around 0.66, comparable with previously published studies, the multiclass framework reached a macro-averaged AUROC of 0.690, with balanced accuracies for individual classes approaching or exceeding the binary models. The strongest performance was observed for Class 0 (AUROC = 0.723, BA = 0.580), the classical case presentation of CMVD. For Class 2, the classic control group, the model performed similarly (AUROC = 0.721), indicating that the model strongly predicted classical case/control presentations. Performance remained robust across less prevalent classes, despite expected challenges associated with class imbalance and reduced sample sizes. Class 1 and Class 3, which represent smaller subsets of the cohort, achieved an AUROC of 0.648 and 0.669, respectively, demonstrating that the model was able to generalize its predictive capacity beyond the dominant classes. Multiclass classification models consistently yielded lower (better) Brier scores than binary models. Results for the MLR model are reported in **Supplemental Table S4**.

Measuring the class proportions across stratified PRS risk tertiles, shown in **Figure 3A**, revealed patterns supporting the clinical relevance of the endotypes. Class 0 showed a clear positive association with high PRS, suggesting that individuals with higher genetic risk are more likely to have the most pathologically consistent endotype. In contrast, Class2 demonstrated a decline in frequency with higher PRS, consistent with its characterization as a low-risk phenotype. Class 1 and Class 3 remained relatively stable across PRS tertiles. These patterns underscore how integrating endotyping with genetic risk stratification can reveal nuanced and biologically grounded subphenotypes that would be obscured in binary classifications. Based on SHAP analysis, shown in **Figure 3B**, each endotype displayed unique proteomic drivers as key features. Class 0, representing the traditional CMVD case phenotype, was distinguished by proteins PAR-1, REN, BOC, and BNP, among others. In Class 2, representing the traditional control group, REN, BNP, and IL1RL2 contributed the most to the classification. Both cases and controls included the PRS as an essential feature, consistent with **Figure 3A**. Class 1 and Class 3 represented the intermediate groups but prioritized different features. Class 1 prioritized FGF-23 and TIE2 and included enrollment age as a key feature. Class 3, in contrast, had biological sex as the most important SHAP feature, followed by SLAMF7, CEACAM8, THBS2, IL-27, and other proteins.

**Fig. 3.**
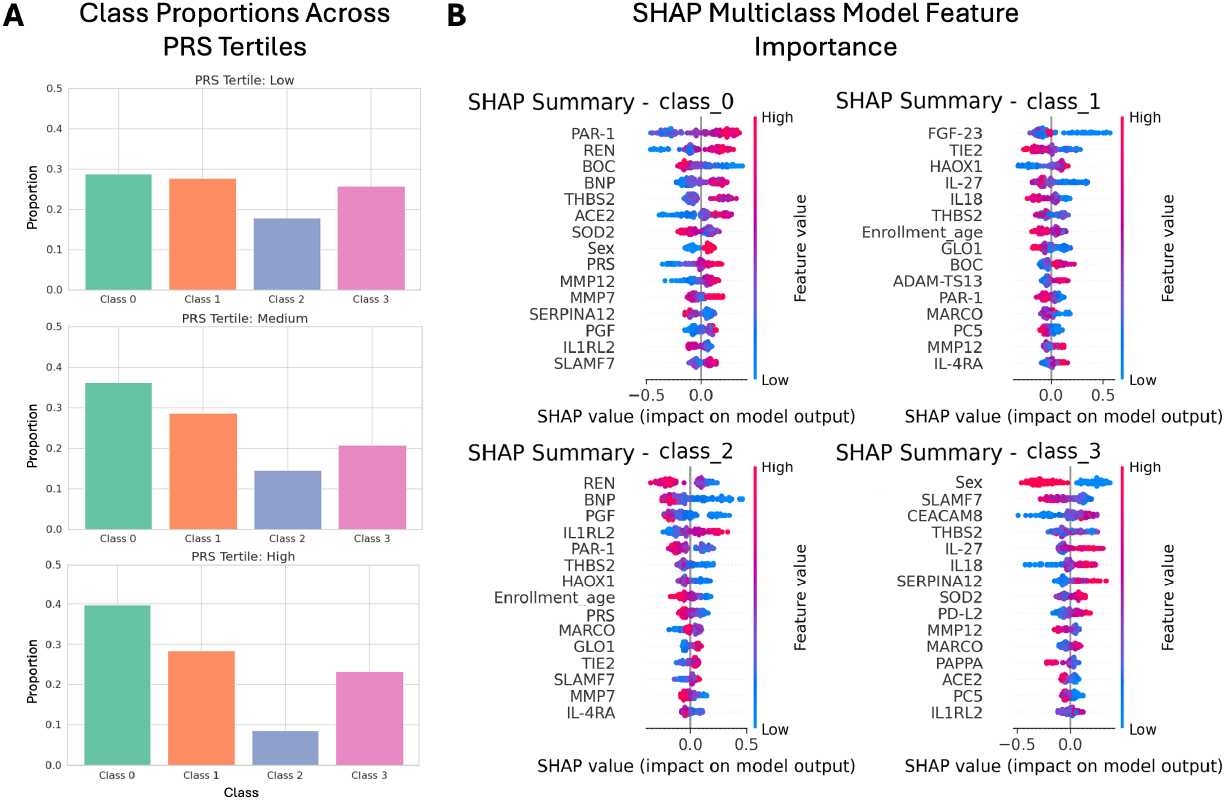
Multiclass model performance. A) Class stratification across PRS tertiles. B) SHAP multiclass feature importance.

## 4. Discussion

### 4.1. Information Gain from Imaging and Proteomics Data

This study represents a novel and comprehensive effort to predict CMVD using multi-modal imaging, proteomics, and genetic data, integrated through machine learning approaches. Unlike existing clinical models, our models were specifically designed to predict risk in earlier, potentially asymptomatic stages^10^. This shift toward earlier identification is critical for a condition like CMVD, which often goes undetected until significant vascular damage has occurred^5,7^. By excluding disease-driven clinical comorbidities, we demonstrated that genetic and proteomic features can drive predictive accuracy. Our models, particularly EN and XGBoost, achieved strong performance across all metrics when incorporating proteomics, surpassing demographic and genetic models alone, and existing clinical models. These improvements are especially meaningful for risk stratification in clinical populations where overt signs of disease have not yet emerged, offering the potential for earlier intervention and monitoring. The relatively poorer performance of the autoencoders and FNNs likely reflects the limited feature space and relatively small sample size in this dataset^36,41^. As sample sizes increase, performance of these methods may improve; however, our results underscore the robustness of linear and tree-based methods for proteomics-guided risk prediction in CMVD.

In addition to predictive gains in modeling, our findings provide biological insights into CMVD pathogenesis. Previously known markers of cardiac stress such as age, BNP, and REN were top contributors to model performance, in line with the previous model in Prescott *et al*. ACE2, another mediator art of the renin-angiotensin-aldosterone system (RAAS), not seen in other clinical models, was significant in our models^10,44–46^. These findings support expert consensus that targeting RAAS may be helpful for CMVD, despite not being fully captured by prior risk models. Additional proteomic signals of interest from top models included ADM, THBS2, and PTX3, among others. ADM is a vasodilatory peptide that helps maintain vascular integrity, while THBS2 and PTX3 have been implicated in vascular inflammation^47–49^. These findings suggest that proteomic profiling may enhance risk stratification and improve our understanding of CMVD pathophysiology.

### 4.2. Image-Derived Endotyping Significantly Improves Risk Prediction

CMVD is a multifactorial disease with several underlying mechanisms. While a MBFR threshold of 2 for PET is often used for binary classifications clinically, this may obscure important heterogeneity in pathophysiology and patient trajectory. By applying multiclass modeling and endotyping approaches, we demonstrate that stratifying patients into biologically-informed subgroups improves risk prediction and reveals distinct associations with genetic and proteomic features. Our results identified four major endotypes, each with unique proteomic signatures and potential clinical implications. The PRS demonstrated stronger risk stratification capabilities across the standard case/control classes, with weaker resolution in the intermediate classes. Class 0 represents the classic CMVD case group, characterized by severely impaired flow and a strong signal of vascular injury and remodeling. Top SHAP features for this group included PAR-1, BNP, BOC, REN, ACE2, and SOD, all of which are implicated in endothelial dysfunction, neurohormonal activation, and oxidative stress^44,46,50,51^. These findings are consistent with known pathophysiological mechanisms of microvascular ischemia and validate Class 0 as the most prototypical disease phenotype^1,8,11^. In contrast, Class 2 represents the classic control group, characterized by preserved MBFR and a biomarker profile indicative of vascular homeostasis. Essential proteins in this group included REN, BNP, PGF, IL1RL2, PAR-1, and THBS2, which, despite some overlap with Class 0, appear with inverse directionality. This group likely reflects a more regulated state, with expression patterns consistent with effective vascular signaling and less inflammation^44,46,49,50,52^. Class 1 is an intermediate phenotype with largely preserved flow but early indications of vascular stress. The top features in this group, FGF-23, TIE2, IL-27, and PRSS27, are linked to dysregulation in vascular tone, angiogenesis, and immune signaling^53–56^. These patients may be at higher risk of progression and warrant closer monitoring. Class 3 presented with notable sex-specific influences, with sex emerging as a top SHAP feature and showing a wide, polarized distribution, indicating strong and variable effects. This is consistent with findings of sex-specific modulation in stress and rest flow^8,57^. The key proteins driving this endotype included SLAMF7, CEACAM8, and SERPINA12, suggesting an immunomodulatory signature potentially influenced by sex hormones or variations in the immune system^58–60^. These patients may represent a CMVD phenotype that is not easily captured by traditional perfusion metrics.

This multiclass endotyping framework outperformed binary models in uncovering distinct biological patterns and enhancing associations with both PRS and proteomic profiles. The AUROC scores across all classes were high, suggesting that the model effectively distinguished between patients at varying levels of CMVD risk. Class 0 and Class 2 achieved AUROCs of 0.723 and 0.721, respectively, while the overall multiclass AUROC reached 0.690, indicating robust ranking capability. However, metrics such as precision and recall were generally lower, particularly for minority classes (Class 2 had a recall of 0.145), largely due to class imbalance introduced by the clustering strategy. Importantly, F1-scores remained relatively high and Brier scores remained low across all classes (0.107-0.197 per class, 0.166 overall), suggesting that while exact class predictions may have been imperfect, predicted probabilities were well-calibrated^61,62^. The results underscore the value of redefining disease states using systems-level data and machine learning tools. This approach enables a more nuanced and interpretable understanding of CMVD heterogeneity, with potential implications for risk stratification, biomarker discovery, and therapeutic targeting.

### 4.3. Future Approaches and Limitations

This study has some limitations and opportunities for improvement. The patient cohort was drawn from a single center, the Hospital of the University of Pennsylvania, and includes only patients referred for perfusion PET imaging as part of routine clinical care. This population thus represents a higher-risk clinical group, which may not be generalizable to the broader population. However, this enrichment for disease enhances the ability to detect biological signals associated with CMVD. Additionally, the proteomic analysis was restricted to angiogenesis-related proteins from the OLINK Cardiovascular II panel, which may not capture other potential mechanisms, such as immune or metabolic pathways. The effect sizes for the PRS were derived from a CAD GWAS, rather than a CMVD-specific GWAS. This was necessary due to the lack of sufficiently powered GWAS on CMVD, but the genetic risk captured may not fully represent the genetic architecture unique to microvascular dysfunction. Future availability of large-scale CMVD GWAS will be critical to improve PRS relevance and specificity for this condition.

This study is the first to integrate imaging-derived measurements with genetics and proteomics to characterize risk and define CMVD endotypes in an unsupervised manner. Furthermore, our use of a multiclass classifier and SHAP-based interpretation provides novel insights into the clinical and molecular distinctions across CMVD subtypes. Our future work will expand proteomic coverage and incorporate additional clinical data, such as comorbidities and laboratory measurements to improve prediction. Another key direction is the further investigation of biological sex differences within CMVD endotypes, particularly in the intermediate group (Class 3), which exhibits early vascular signaling patterns that may present differently across sexes. Overall, this work supports the continued development of omics integration and endotyping to guide personalized risk assessment.

## Data Availability

All summary data will be made available, and the supplemental tables include full analyses results.

https://ritchielab.org/publications/supplementary-data/psb-2026/cmvdriskpred

## 5. Acknowledgements

We acknowledge the Penn Medicine BioBank and Regeneron Genetics Center for providing data and thank the patient-participants of Penn Medicine who consented to participate in this research program. The PMBB is approved under IRB protocol# 813913 and supported by the Perelman School of Medicine at the University of Pennsylvania, a gift from the Smilow family, and the National Center for Advancing Translational Sciences of the National Institutes of Health under CTSA award number UL1TR001878.

## 6. Appendix

Supplemental figures and tables are available at: https://ritchielab.org/publications/supplementary-data/psb-2026/cmvdriskpred

All modeling and analysis code are available at: https://github.com/rvenkatesh99/Proteomics_Modeling

