## Supplemental Figures S1-5 for "Integrating Imaging-Derived Clinical Endotypes with Plasma Proteomics and External Polygenic Risk Scores Enhances Coronary Microvascular Disease Risk Prediction^†^"

#### Supplemental Figure S1: Distribution of Image-Derived Flow Measurements

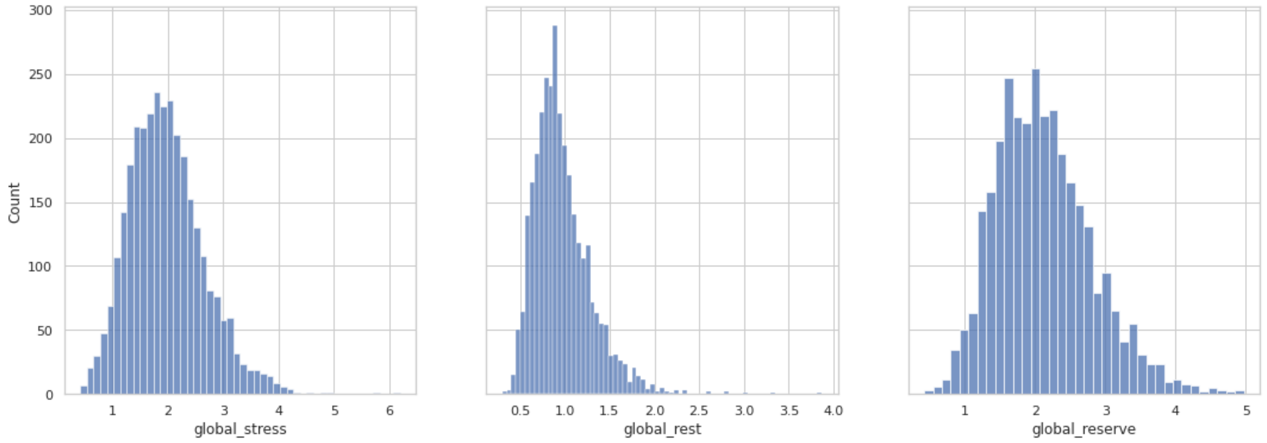


**Supplemental Fig. S1. Distribution of image-derived flow measurements. Histograms depict the distribution of stress and rest flow, as well as flow reserve; quantified as the ratio between stress and rest.**

### Supplemental Figure S2: SHAP Summary Plots for Model 3

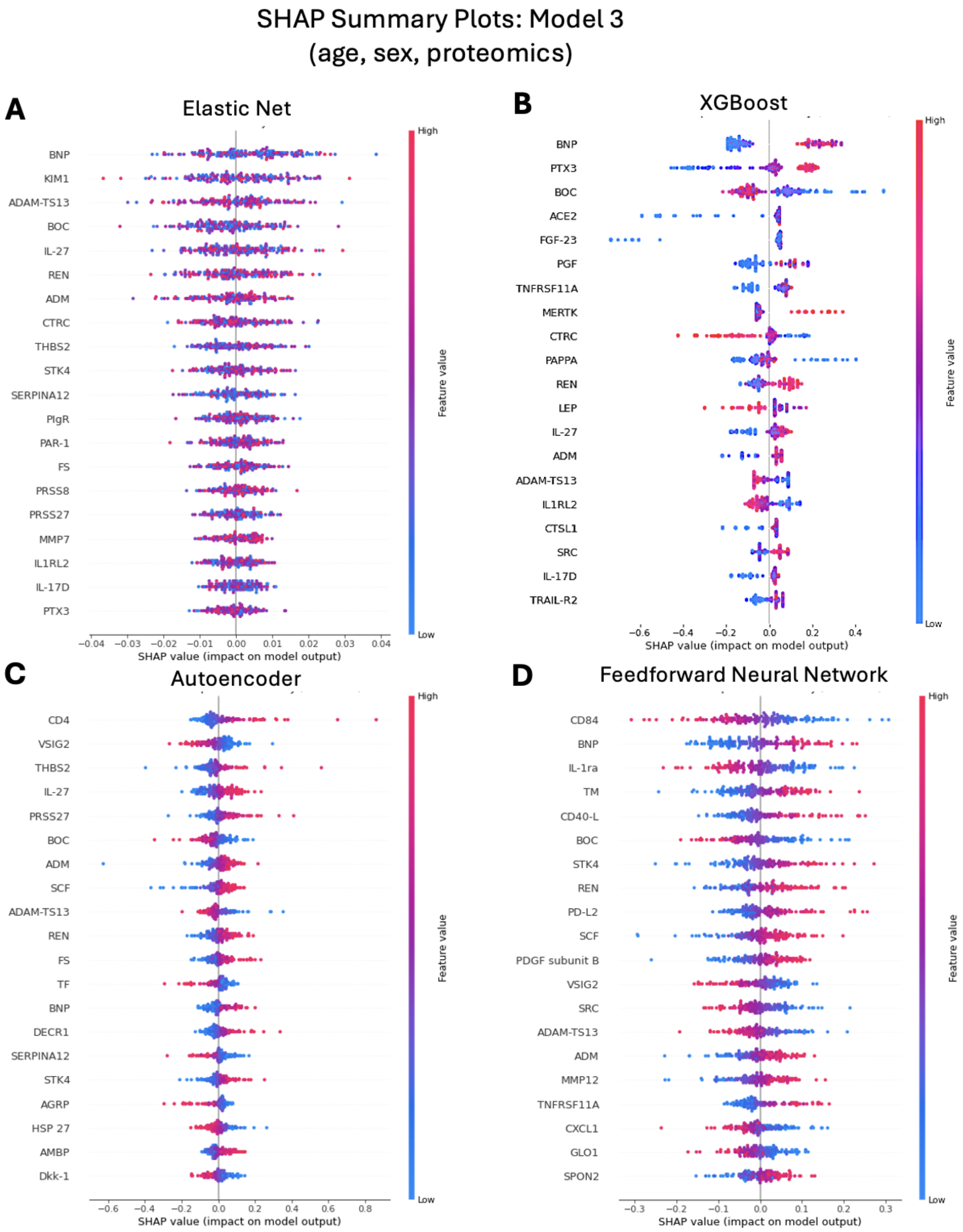


**Supplemental Fig. S2. SHAP Summary Plots for Model 3 (age, sex, proteomics) depict the top 15 features across each model, ranked by their overall impact on model output. Feature importance is visualized by the magnitude and direction of SHAP values, with color indicating the feature value (high - red or low - blue).**

#### Supplemental Figure S3: SHAP Summary Plots for Model 4

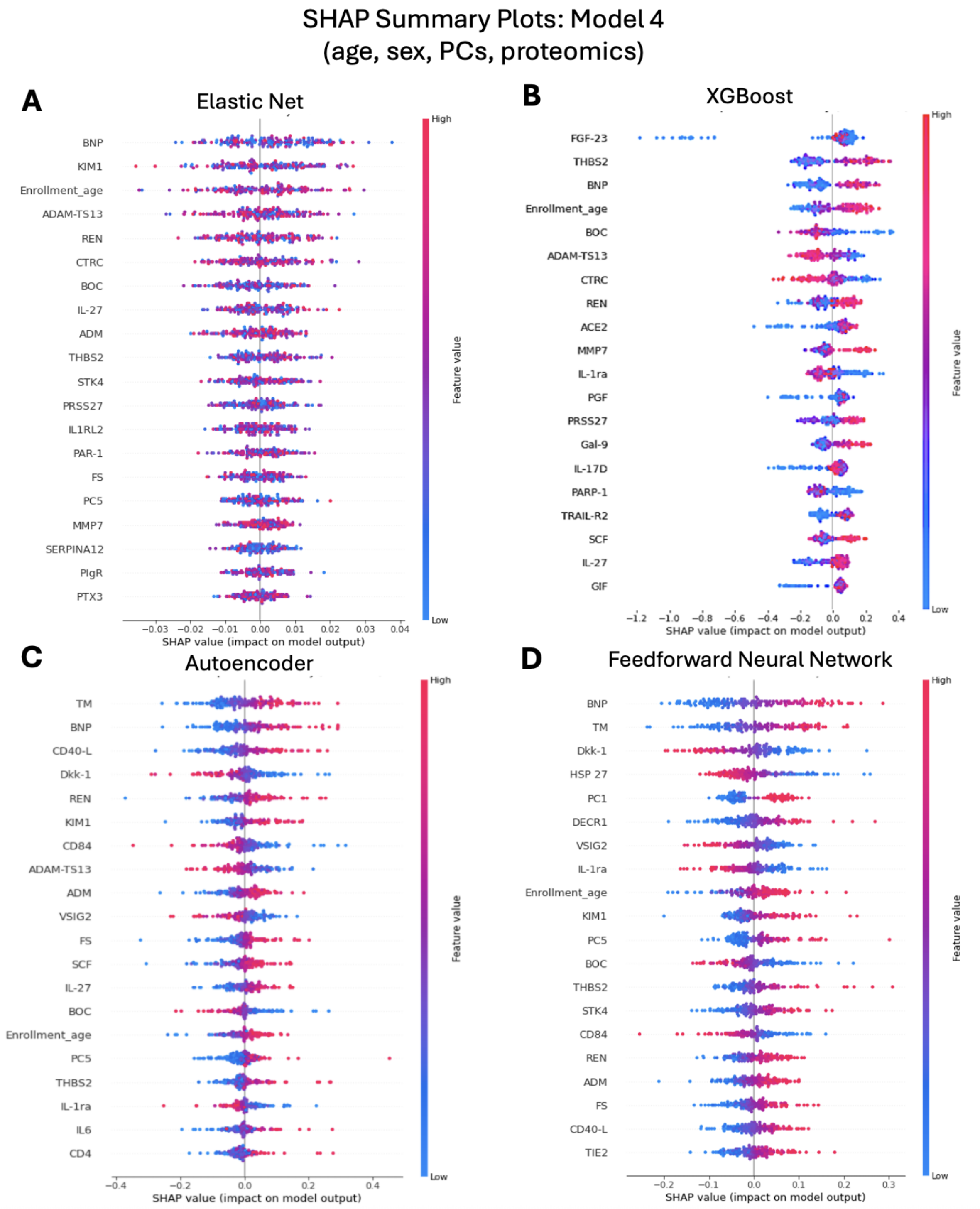


**Supplemental Fig. S3. SHAP Summary Plots for Model 4 (age, sex, PCs 1-5, proteomics) depict the top 15 features across each model, ranked by their overall impact on model output. Feature importance is visualized by the magnitude and direction of SHAP values, with color indicating the feature value (high - red or low - blue).**

#### Supplemental Figure S4: SHAP Summary Plots for Model 8

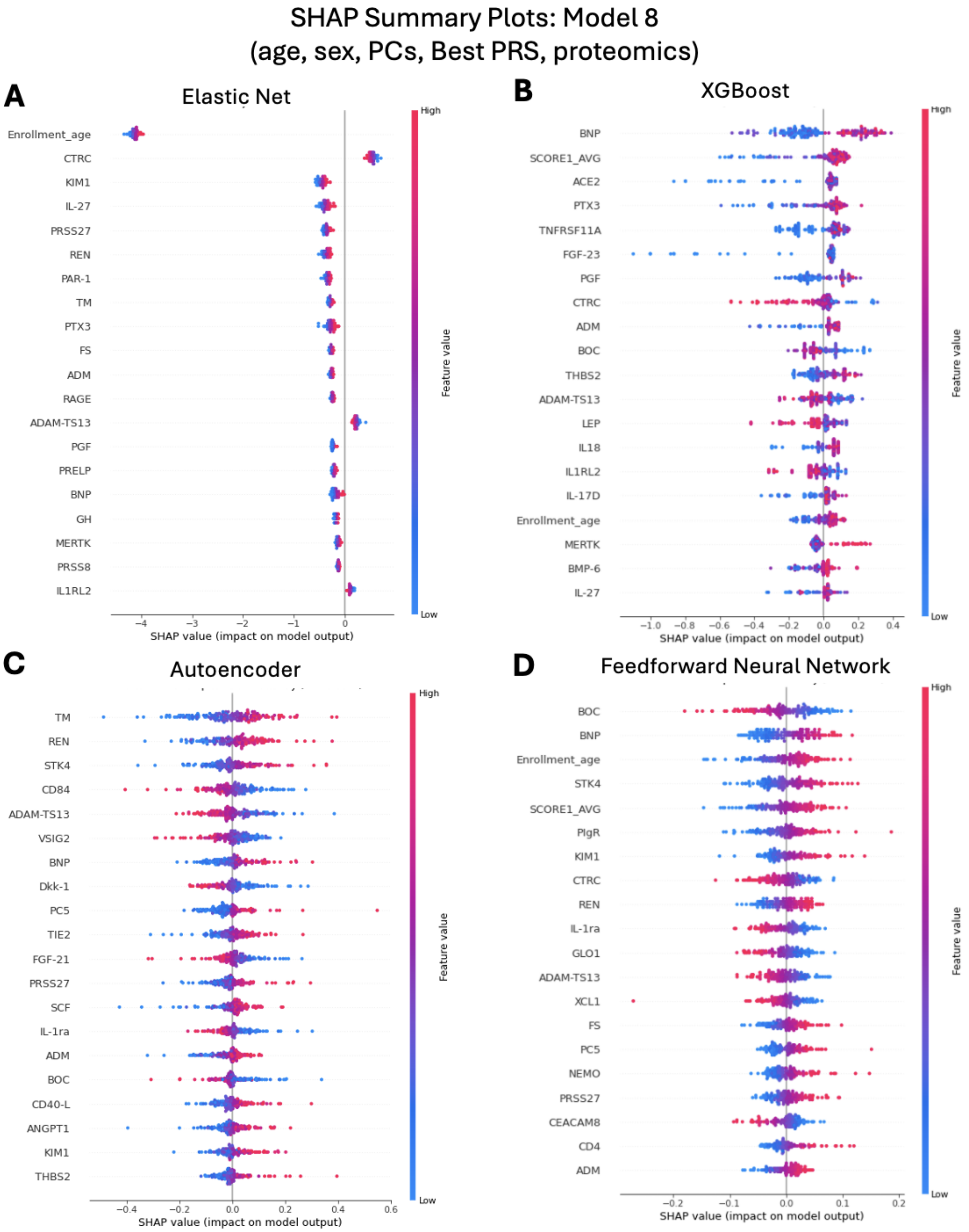


**Supplemental Fig. S4. SHAP Summary Plots for Model 8 (age, sex, PCs 1-5, CMVD Targeted PRS, proteomics) depict the top 15 features across each model, ranked by their overall impact on model output. Feature importance is visualized by the magnitude and direction of SHAP values, with color indicating the feature value (high - red or low - blue).**

#### Supplemental Figure S5: Elbow Plot for Clustering

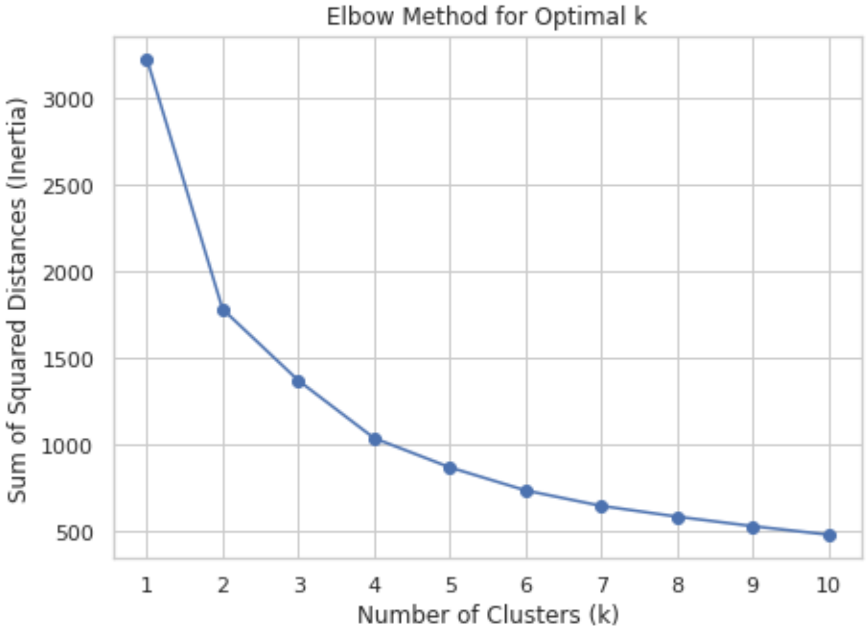


**Supplemental Fig. S5. Elbow plot for determining optimal number of clusters. The elbow of the curve occurs at 4, so 4 clusters were determined to be optimal.**
